# Regional variation in cardiovascular genes enables a tractable genome editing strategy

**DOI:** 10.1101/2022.03.03.22271786

**Authors:** Vikki A. Krysov, Rachel H. Wilson, Nicholas S. Ten, Nathan Youlton, Hannah N. De Jong, Shirley Sutton, Yong Huang, Chloe Reuter, Megan E. Grove, Matthew T. Wheeler, Euan A. Ashley, Victoria N. Parikh

## Abstract

Recent rise in the number and therapeutic potential of genome engineering technologies has generated excitement in cardiovascular genetics. One significant barrier to their implementation is costly and time consuming reagent development for novel unique variants. Prior data have illuminated the potential for regional clustering of disease-causing genetic variants in known and potential novel functional protein domains. We hypothesized that most cardiovascular disease-relevant genes in ClinVar would display regional variant clustering, and that multiple variants within a regional hotspot could be targeted with limited prime editing reagents. We collated 2,471 high confidence pathogenic or likely pathogenic (P/LP) missense and truncating variants from 111 cardiovascular disease genes. We then defined a regional clustering index (RCI), the percent of P/LP variants in a given gene located within a pre-specified window around each variant. At a window size commensurate with maximally reported prime editing extension length, 78bp, we found that missense variants displayed a higher mean RCI than truncating variants. We next identified genes particularly attractive for pathogenic hotspot-targeted prime editing with at least 20 P/LP variants and observed that the mean RCI in missense variants remained higher than for truncating variants and for rare variants observed in the same genes in gnomAD (Mean±SD RCI_78bp_: P/LP ClinVar Missense=5.2±5.9%; P/LP ClinVar Truncating=2.1±3.2%, gnomAD Missense=2±3.2%, gnomAD Truncating=1.1±2.4%. p<2.2e-16; ANOVA with post hoc Bonferroni correction). Further, we tested the feasibility of prime editing for multiple variants in a single hotspot in KCNH2, a gene with a high mean missense variant RCI. Sixty variants were induced in this hotspot in HEK293 cells with CRISPR-X. Correction of these variants was attempted with prime editing using two overlapping prime editing guide RNA extensions. The mean prime editing efficiency across CRISPR-X-enriched variants within this hotspot was 57±27%, including 3 P/LP variants. These data underscore the utility of pathogenic variant hotspots in the diagnosis and treatment of inherited cardiovascular disease.

---

Recent rise in the number and therapeutic potential of genome engineering technologies has generated excitement in cardiovascular genetics. One significant barrier to their implementation is costly and time consuming reagent development for novel unique variants. We have previously shown that disease-associated variants cluster in functional protein domains where variants in the general population are lower frequency.^1,2^ These findings may provide an opportunity to pre-emptively target multiple pathogenic variant clusters (“pathogenic hotspots”) with pre-designed reagents. Prime editing, a search-and-replace editing technology, can overwrite genomic sequences up to 78 base pairs in length with a reverse transcriptase guided by a Cas9 nickase and prime editing guide (peg)RNA.^3^ We hypothesized that most cardiovascular disease-relevant genes in ClinVar would display regional variant clustering, and that multiple variants within a regional hotspot could be targeted with limited prime editing reagents.

We collated 111 cardiovascular disease genes in ClinVar. From these, missense or truncating (stop-gain or frameshift) pathogenic and likely pathogenic (P/LP) variants were included. These were restricted to those with multiple depositors in agreement regarding pathogenicity. Genes with only one high-confidence variant were then excluded, yielding 2,471 variants. To quantify variant clustering, we examined the number of neighboring P/LP neighbors for each P/LP variant within windows ranging from 10 to 200bp. We found that P/LP missense variants have more P/LP missense neighbors than P/LP truncating variants have P/LP truncating neighbors (Figure 1A). To determine the number of prime editing reagents needed to address pathogenic hotspots across entire genes, we defined a regional clustering index (RCI): the percent P/LP variants in a given gene located within a pre-specified window around each variant. At a window size commensurate with maximally reported prime editing extension length (78 bp), mean RCI was 9.3±15.3% for P/LP missense variants versus 2.8±6.4% for P/LP truncating variants (Mean±SD; p <2.2e-16 (student’s T-test), Figure 1B). Therefore, in the average cardiovascular disease gene, it is possible that 11 or fewer pegRNAs can address all known pathogenic variants.

**Figure 1.**
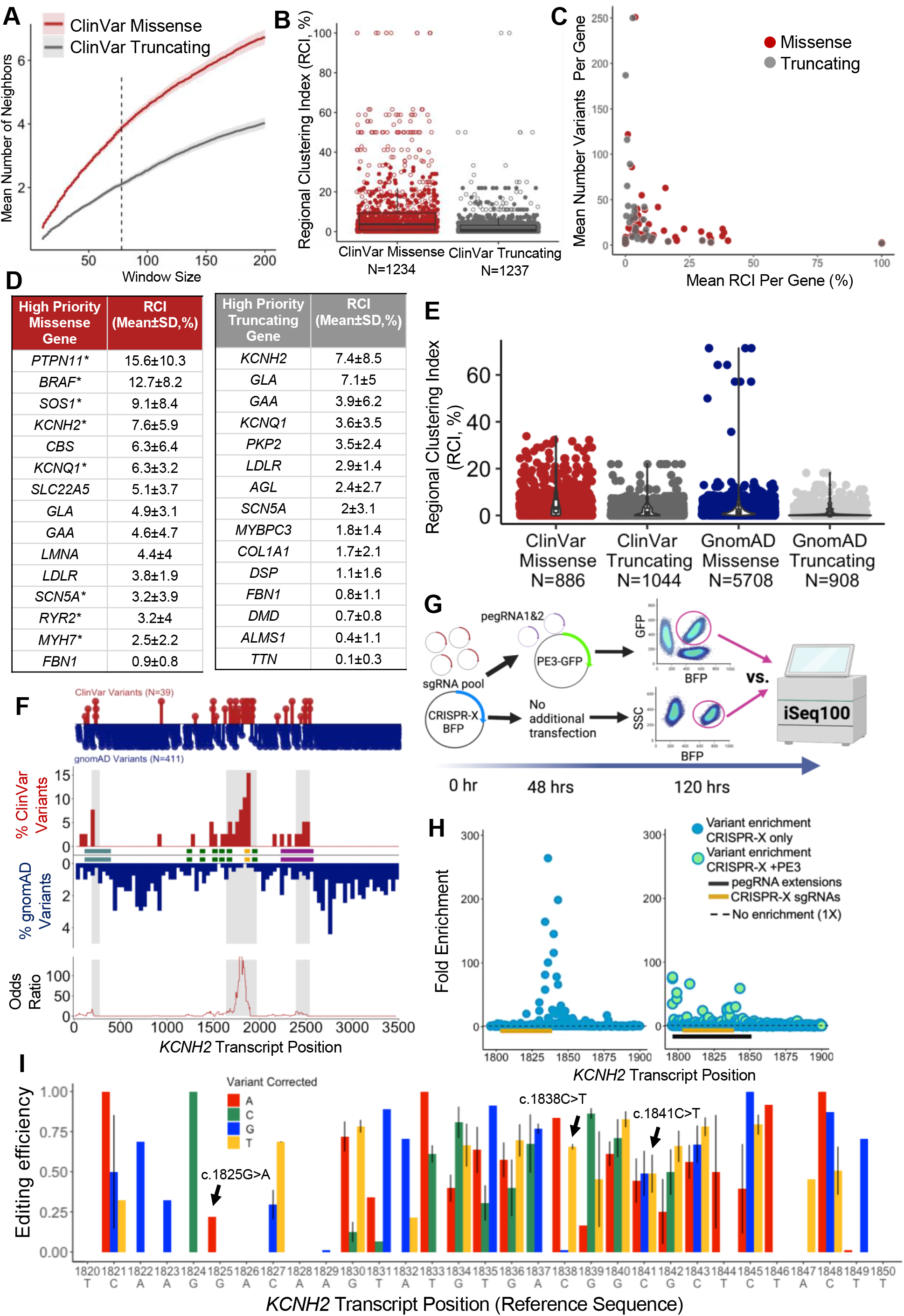
Disease-associated missense variants cluster in functional hotspots and can be targeted using prime editing technology, as demonstrated by the KCNH2 gene. (**A**) Mean number of P/LP neighbors for missense variants (red) and truncating variants (grey) within windows from 10-200bp around each variant (Mean±SE, N=1234 P/LP missense variants and N=1237 P/LP truncating variants). (**B**) Per-variant RCI for a 78 bp window in P/LP missense variants and P/LP truncating variants. Filled points reflect variants in high priority genes with >20 P/LP missense or truncating ClinVar variants. (Mean±SD RCI_78bp_: P/LP ClinVar Missense =9.3±15.3%; P/LP ClinVar Truncating=2.8±6.4%; p <2.2e-16 by student’s T-test). **(C)** Distribution of mean RCI and number of variants per gene illustrating outliers with <20 P/LP missense (red) or truncating (grey) variants. **(D)** High priority CV disease genes (≥20 P/LP variants) for hotspot directed genome editing. * indicates reported gain of function mechanism (not exclusive of loss of function mechanism in other variants). **(E)** High priority P/LP missense variants from ClinVar have a higher RCI than rare population missense variants in the same genes (p < 2.2e-16), and the same is true for P/LP versus population truncating variants (p= 8.0e-9). (Mean±SD RCI_78bp_: P/LP ClinVar Missense=5.2±5.9%; P/LP ClinVar Truncating=2.1±3.2%, gnomAD Missense=2±3.2%, gnomAD Truncating=1.1±2.4%. p<2.2e-16; ANOVA with post hoc Bonferroni correction). **(F)** Sliding window analysis of *KCNH2* missense variants in ClinVar (red) versus rare variants in gnomAD (blue, *KCNH2* transcript: ENST00000262186.10). Regions with statistical enrichment for pathogenic variation are shaded in grey (FDR 0.1) and overlap with multiple functional domains annotated as shaded boxes along the x-axis. Functional domains labeled on x-axis: PAS domain (light blue), S1-S6 transmembrane domains (green), H5 pore-forming domain (gold), and cyclic nucleotide binding domain (purple). **(G)** Experimental design for pegRNA hotspot targeting in *KCHN2*. **(H)** Fold enrichment of variant frequency in CRISPR-X treated HEK293 cells vs untreated (left panel) and CRISPR-X plus PE3 vs untreated controls. **(I)** Editing efficiency by PE3 for variants statistically enriched in CRISPR-X treated cells (1-(Fold change _CRISPR-X+PE3_/Fold change_CRISPR-X only_)). Error bars indicate standard deviation across 2-4 replicates for variants reaching statistically significant fold increase in the CRISPR-X only group.

We next identified genes particularly attractive for pathogenic hotspot-targeted prime editing. The distribution of mean variant RCI versus number of variants in each gene reveals a predominance of genes with <20 known P/LP variants among those with mean RCI >25% (Figure 1C). In genes with at least 20 P/LP missense variants (high priority genes), the mean RCI in missense variants remained higher than for truncating variants, and rare variants observed in the same genes in gnomAD showed reduced RCI compared to Clinvar P/LP variants (MAF <0.0001, Figure 1D&E). Over half of high priority missense variant genes have been associated with dominant negative or gain-of-function disease mechanisms (Figure 1D). These data are consistent with P/LP missense variant location in known and potentially novel functional transcript regions compared to truncating variants, and indicate that gain-of-function disease mechanisms are frequent in genes with highly clustered P/LP missense variants.

We next tested the feasibility of prime editing for multiple variants in a single hotspot in *KCNH2*, a cause of long and short QT syndrome with a high mean missense variant RCI. We used a 78bp sliding window approach to identify regions of *KCNH2* enriched for P/LP missense variants versus population-associated variants (gnomAD, MAF<0.0001) as we have previously described.^1^ Windows significantly enriched for P/LP variants in the *KCNH2* transcript aligned with known functional domains (FDR 0.1,^1^ Figure 1F).^4^ Positions c.1800-1850 showed significant P/LP enrichment, and overlap the H5 intramembrane pore-forming domain.

We first introduced multiple variants in this hotspot in HEK293 cells using CRISPR-X, a targeted hyper-mutagenesis tool that creates mutations within 50 bp of guideRNAs.^5^ This was followed by prime editing with generation PE3 including 2 pegRNAs with extensions covering the length of the hotspot,^3^ and FACS for editing enrichment (Figure 1G, detailed methods in supplement). Resultant editing efficiency was quantified by comparing variant prevalence in CRISPR-X+PE3 vs. CRISPR-X alone (iSeq100). CRISPR-X alone showed up to ∼300-fold increase per variant compared to untreated cells, localized at *KCNH2* c.1820-1850 (Figure 1H, blue, 66% of variants significantly enriched (FDR 0.05)). In CRISPR-X+PE3, a much smaller fold increase was observed (Figure 1H, blue/green). Nine variants displayed *increased* prevalence in the CRISPR-X+PE3 condition compared to CRISPR-X alone in at least 1 of 4 replicates, likely attributable to persistent Cas9-AID expression during and after PE3 editing. To provide a conservative estimate of prime editing efficiency for these variants, these observations were encoded as 0 editing efficiency. Accounting for this, the mean prime editing efficiency across CRISPR-X-enriched variants within this hotspot was 57±27%, including 3 P/LP variants (Figure 1I, black arrows).

These data underscore the utility of pathogenic variant hotspots in the diagnosis and treatment of inherited cardiovascular disease. That missense P/LP variants in particular cluster in these hotspots informs incorporation of clinical genetic data into disease-specific investigations and also highlights the potential of these hotspots for uniquely-targeted editing approaches. We find that genes with high mean missense RCI are often associated with gain of function disease mechanisms for which existing therapeutic approaches (e.g. gene replacement) will not always be effective, making the hotspot-targeted prime editing we present an attractive solution. Lastly, novel P/LP variants are likely to be identified within functional hotspots, including those identified here, making hotspot-targeted prime editing even more tractable as a prospective therapeutic strategy.

## Data Availability

All data produced in the present study are available upon reasonable request to the authors

## Acknowledgments

This work was funded by NHLBI K08HL14318503 and R01HL14484302, the Sarnoff Cardiovascular Research Foundation, the John Taylor Babbitt Foundation, and the Stanford School of Medicine Medical Scholars Fund. We would like to thank Hannah Wand, PhD, Hannah Ison, MS, Mitchel Pariani, MS, Greg Newby PhD, David R. Liu PhD, Gaelen Hess, PhD and Michael Bassik, PhD for their advice regarding study design and genome engineering implementation.

## Supplemental Methods

### ClinVar and gnomAD data compilation

A VCF file including all variants compiled in ClinVar (https://ftp.ncbi.nlm.nih.gov/pub/clinvar/) was obtained on July 13th, 2020. GRCh38/hg38 coordinates were used. We selected variants found within 111 cardiovascular disease genes as identified by expert cardiovascular genetic counselors. All pathogenic/likely pathogenic variants with a 2+ star rating (high-confidence variant) within ClinVar were included. From the missense and truncating variants that were examined, 38 genes with only one high-confidence variant were excluded for further analysis given the inability to describe regional clustering in this group. Population variants were acquired from the Genome Aggregation Database (gnomAD v3.1.2) and were filtered for a minor allele frequency < 0.0001.

### Determination of number of neighbors, Regional Clustering Index (RCI) and sliding window analysis

The number of P/LP neighbors for each variant was determined by calculating the number of missense or truncating variants situated within a certain base pair window (10-200bp total) of that P/LP missense or truncating variant using custom code (available upon request). Regional clustering index (RCI) was calculated by dividing the number of P/LP neighbors for a particular variant by the total number of missense or truncating variants within that gene minus the selected variant and multiplied by 100. SnpEff software was used to annotate the transcript position of the ClinVar KCNH2 missense variants. A Fisher’s exact test was used for the sliding window analysis as described previously ^1^ and modified to recalculate enrichment for 78 bp windows sliding down the transcript by one base pair at a time. Functional *KCNH2* domains were collated from UniProt.^4^

### Guide RNA and pegRNA design and preparation

Guides (sgRNAs) for CRISPR-X were designed using CHOPCHOP version 3 (Labun et al., 2019) to target KCNH2. Guides were ordered as single stranded DNA oligos (IDT, Coralville, IA, USA) then annealed and cloned into pBlueScript using Golden Gate Cloning (New England Biolabs, Ispwich, MA, USA). The pegRNAs were designed using PrimeDesign (Hsu et al., 2021) and ordered as single stranded DNA oligos (IDT, Coralville, IA, USA) and cloned into pU6-pegRNA-GG-acceptor (addgene #132777) as previously described (Anazole et al., 2019). Guides and pegRNAs were prepared for transfection using endotoxin free NucleoBond Midi kit (Macherey-Nagel, Duren, Germany).

### Transfection

The HEK293 cells are grown to 90% confluency then transfected with the CRISPR-X plasmid and guide RNA pool using Lipofectamine 3000 reagents according to manufacturer’s protocol (Thermo Fisher, Waltham, MA, USA). Cells were transfected with 2.0 ug of CRISPR-X and 0.5 ug of the sgRNA pool with 7.5 uL Lipofectamine and 5 uL of P3000 reagent in OptiMEM reduced serum media (Thermo Fisher, Waltham, MA, USA) per well (10^6^ cells per well). The sgRNA pool consists of six guides cloned into pBlueScript plasmid. Cells were examined using fluorescent microscopy 24 hours post transfection for blue fluorescent protein (BFP) then split at a ratio of 2:1 and grown for 48 hours. The cells were then transfected with 1.67 ug of Prime Editing plasmid (PE2-GFP, addgene #132776), 0.55 ug pegRNA, and 0.166 ug of a nicking guide cloned into pBlueScript using the previously described Lipofectamine reagents and conditions. Cells recovered for 48 hours then were sorted for BFP+ and GFP+ using fluorescent activated cell sorting on the Falstaff BD Aria II (BD Biosciences, San Jose, CA, USA). Cells positive for BFP only were collected as the CRISPR-X only sample. Cells positive for BFP and GFP were collected as the prime edited cells. Following FACS, collected cells were centrifuged at 1000xg for 5 minutes and the cell pellet was frozen overnight at −80°C.

### Genomic isolation, library preparation, and sequencing

The cell pellet was thawed and genomic DNA was extracted using 200 uL of Quick Extract (Lucigen, Middleton, WI, USA) according to manufacturer’s protocol. The target region of KCNH2 was amplified using gene specific oligos and Q5 high fidelity polymerase (New England Biolabs, Ipswich, MA, USA). The PCR products were run on a 2% agarose gel and extracted using Qiagen QIAquick Gel Extraction Kit (Hilden, Germany). Following purification, samples underwent preparation for sequencing using the Nextera XT v2 library preparation kit (Illumina, San Diego, CA, USA) according to manufacturer’s protocol and run on the iSeq (Illumina, San Diego, CA, USA).

### Variant fold enrichment and editing efficiency

Data analysis of resultant FASTQ files was performed on a custom pipeline (https://github.com/hndejong/CRISPR-X). Per-variant enrichment and editing efficiency were determined from the resultant counts files with custom R code available upon request. Statistical significance of variant enrichment versus untreated samples was determined by a two proportions test at significance FDR 0.05. Editing efficiency was calculated as (1-(Fold change _CRISPR-X+PE3_/Fold change_CRISPR-X only_)) and standard deviation was calculated across 2-4 experimental replicates for all variants reaching statistically significant fold increase with CRISPR-X treatment alone within a given replicate.

### Construct and Primer Sequences

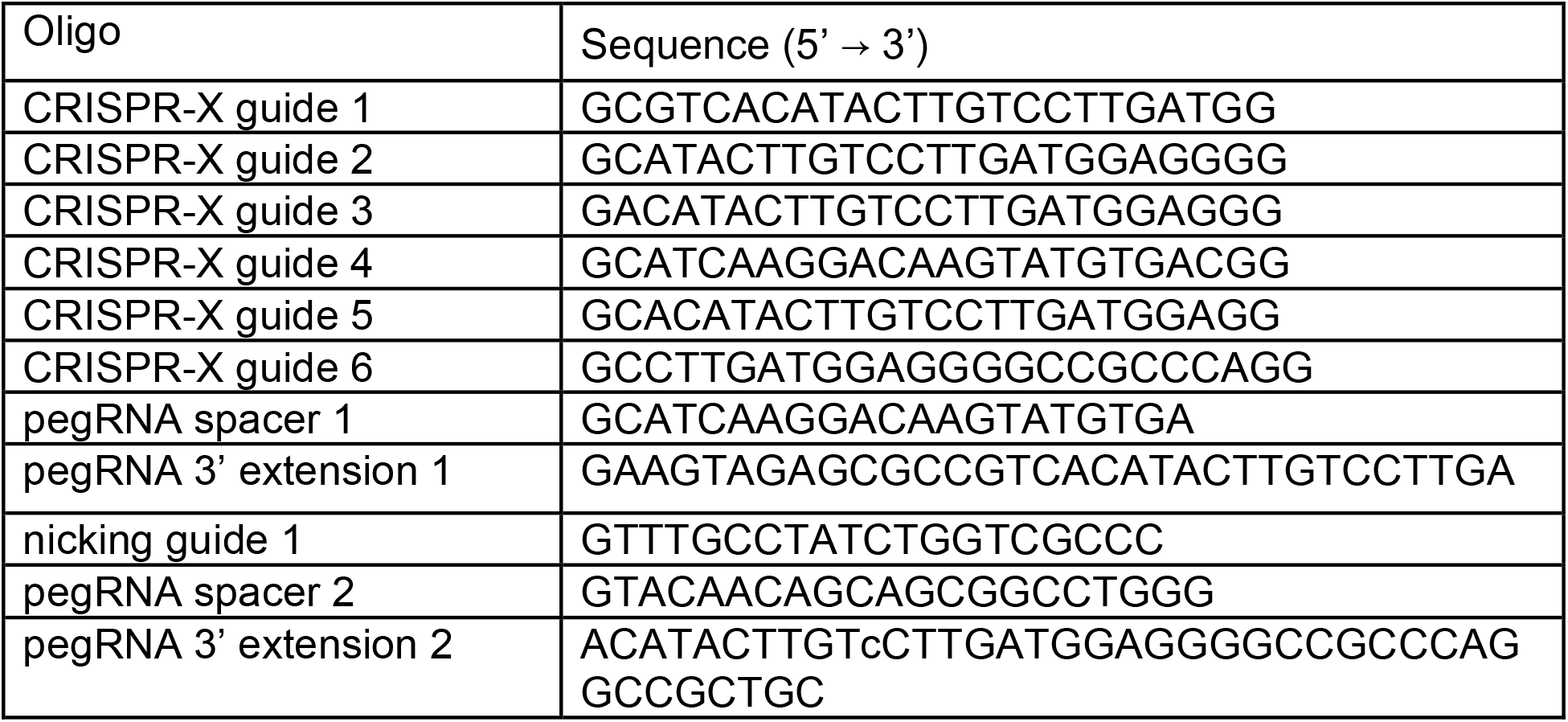
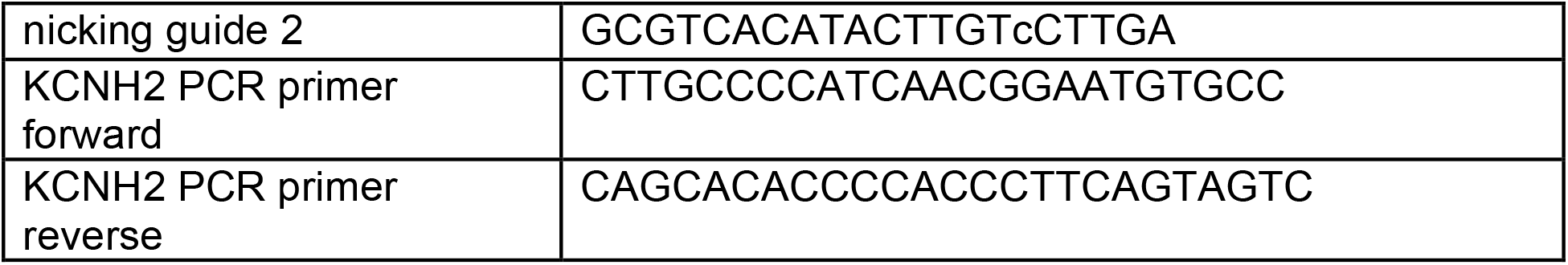

### CRISPR-X construct sequence

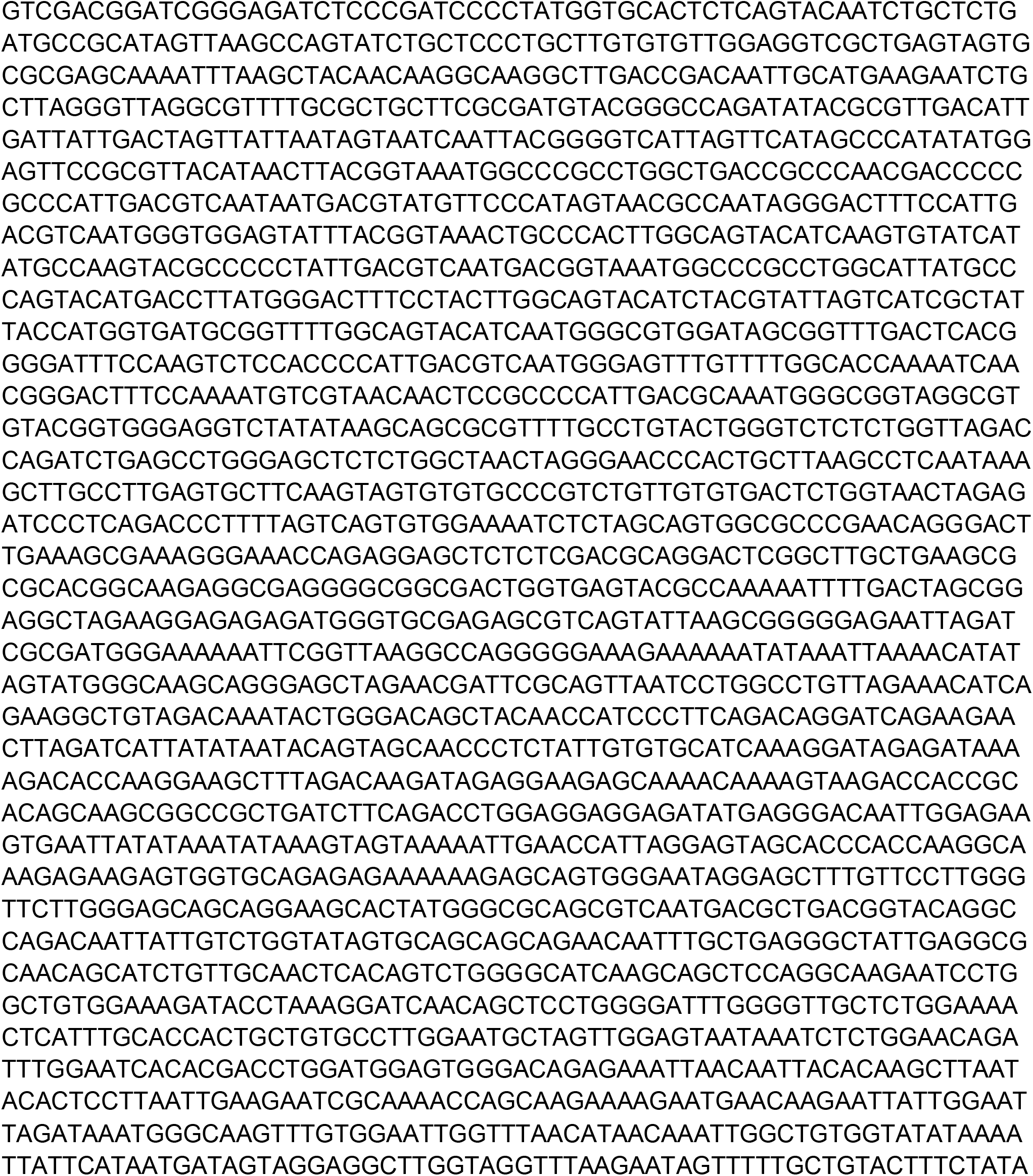

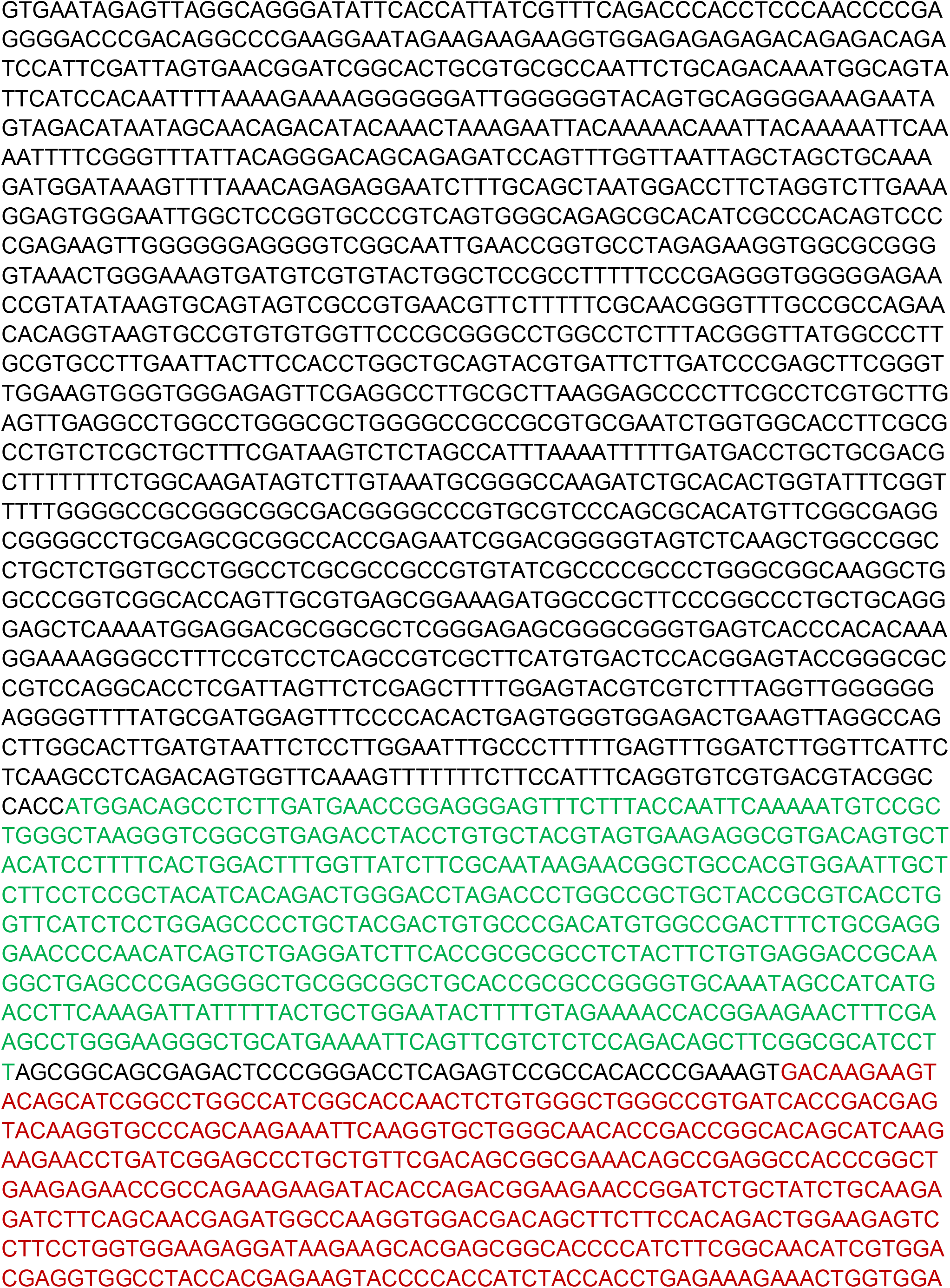

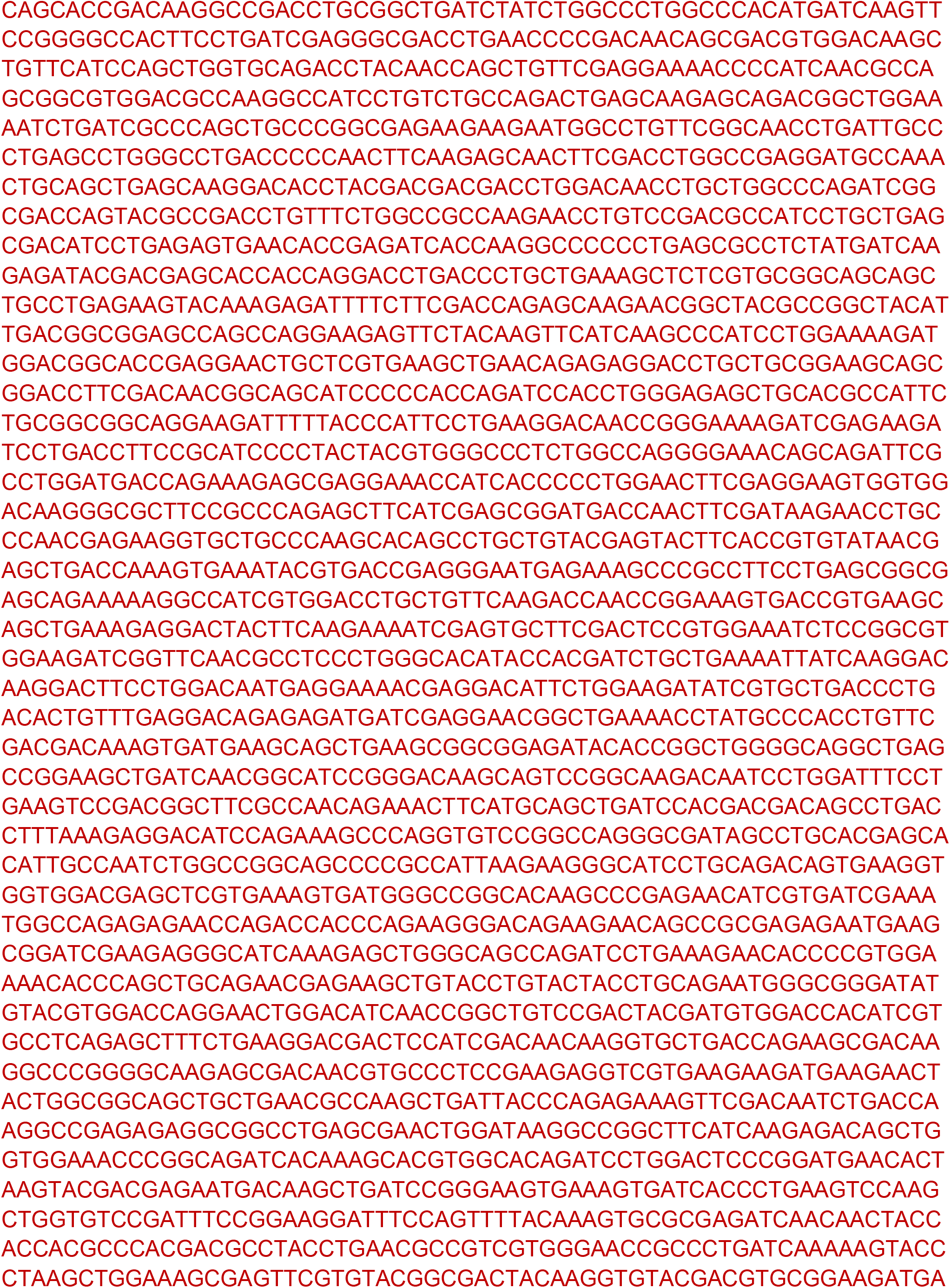

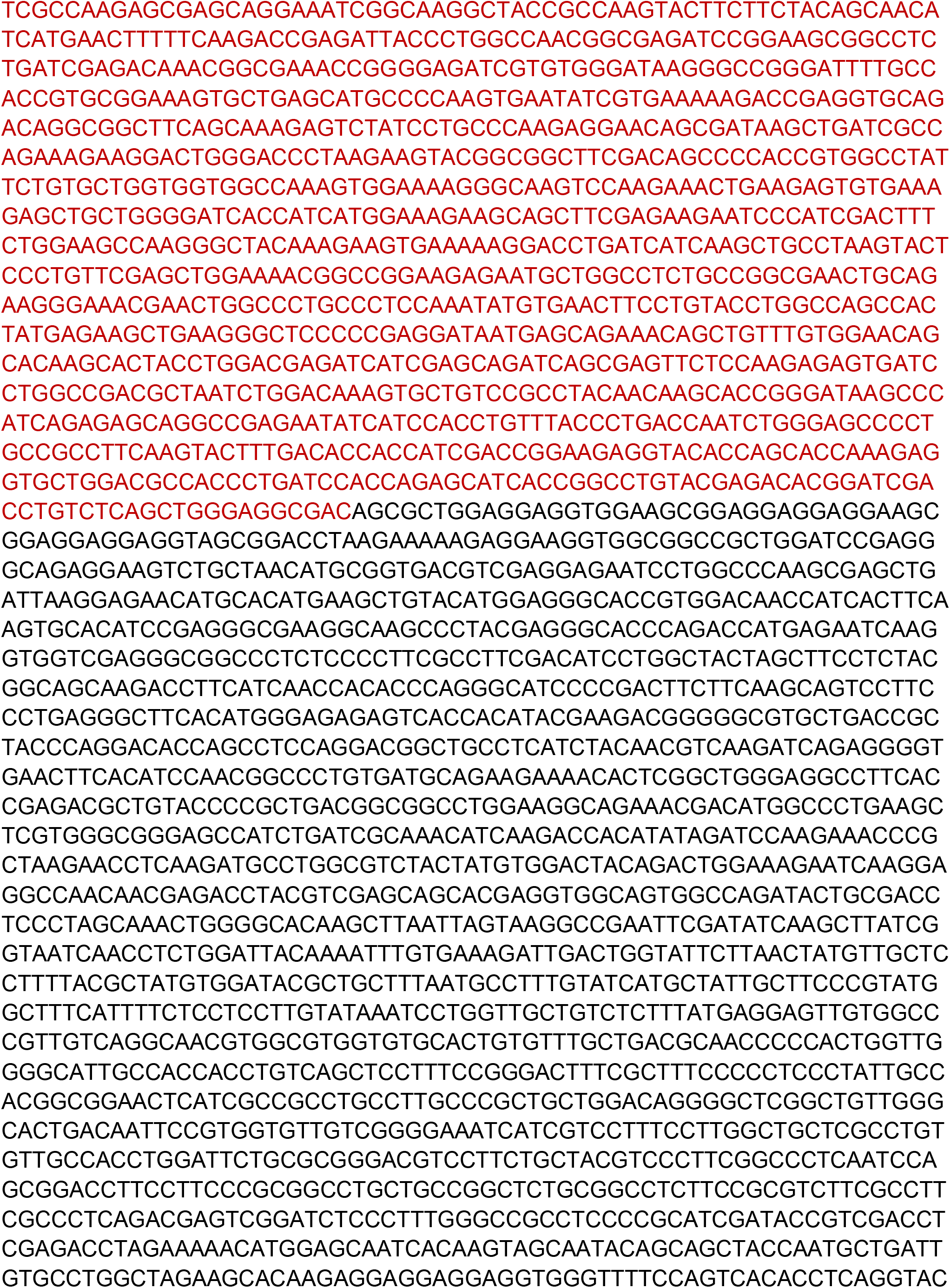

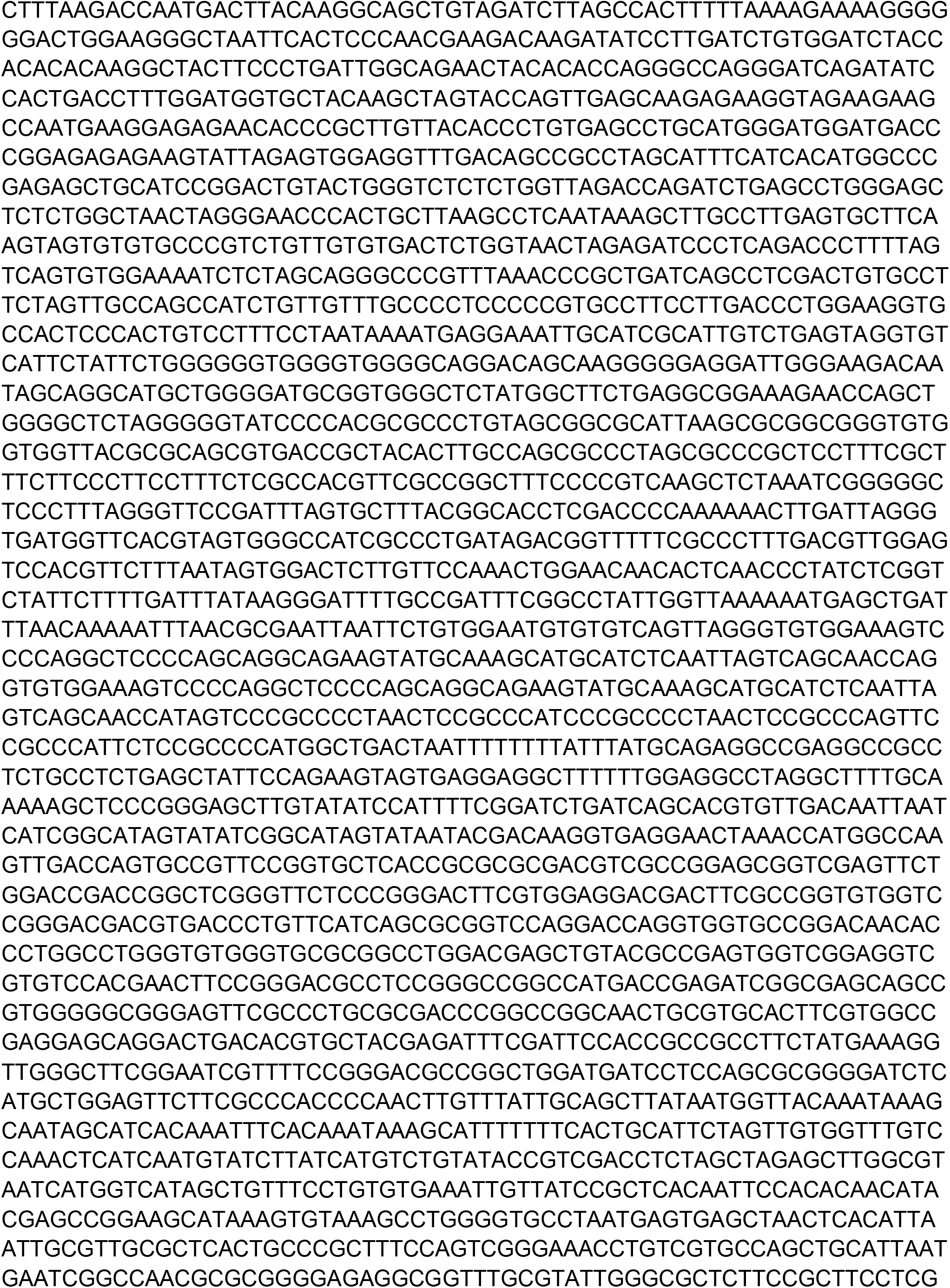

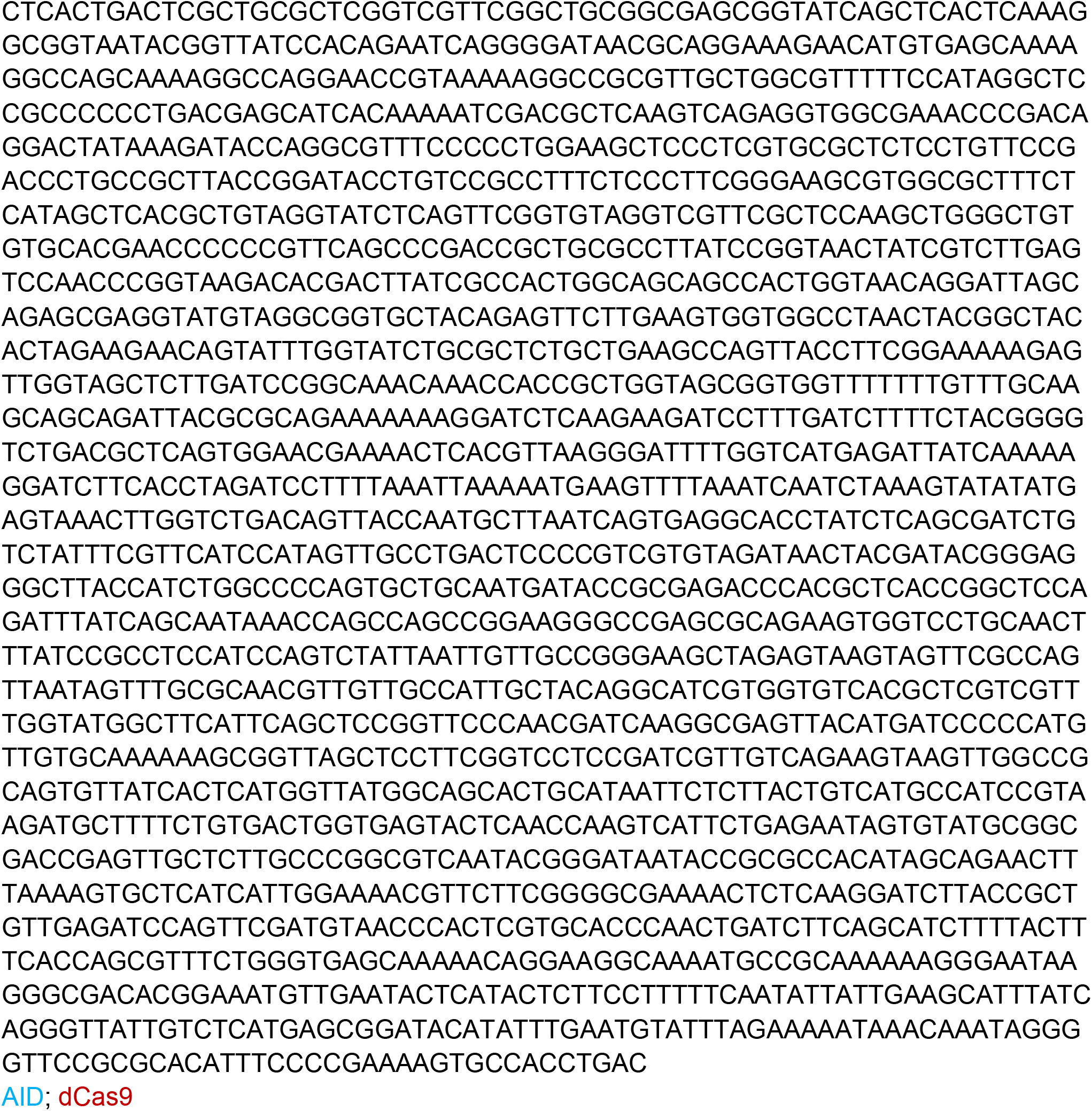

### pBlueScript plasmid sequence

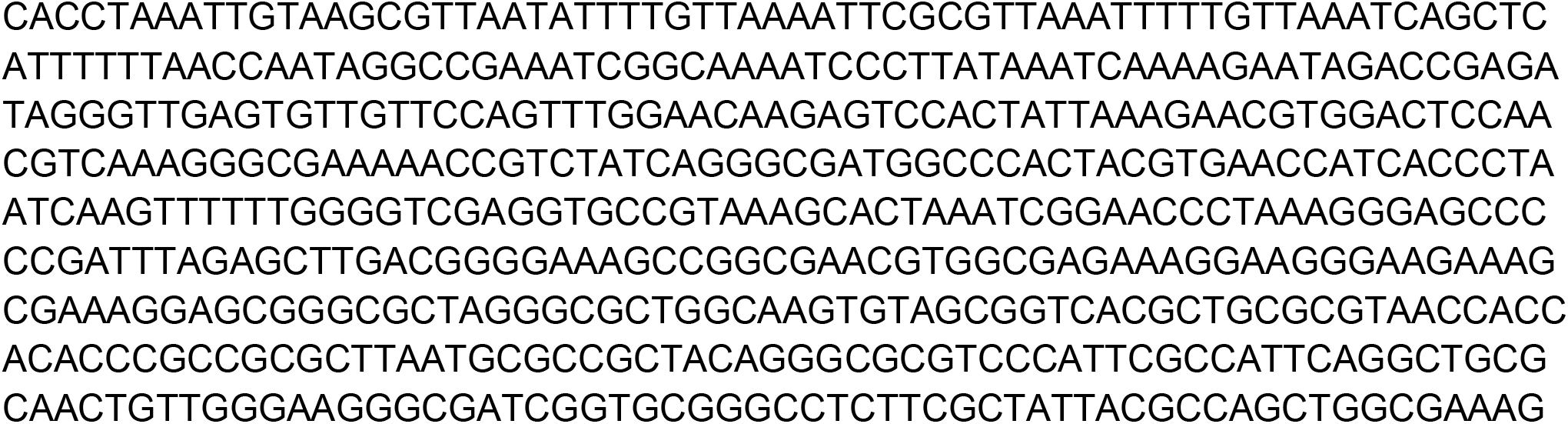

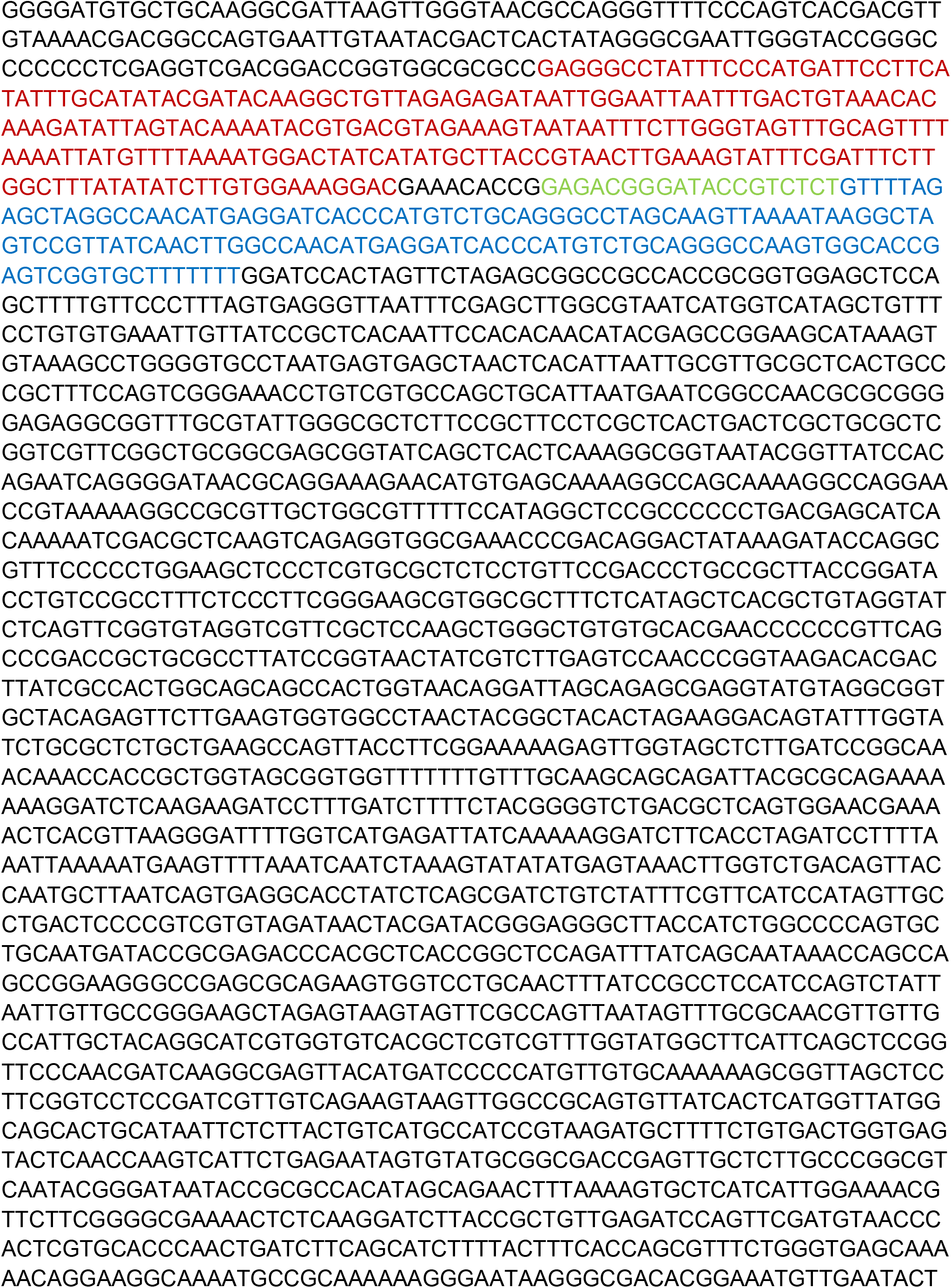

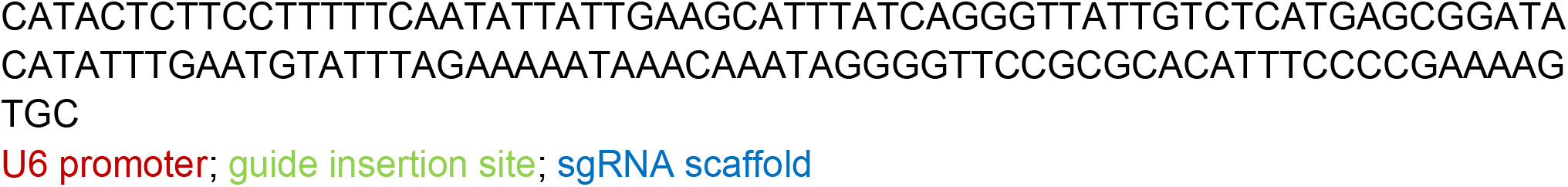

## References

1. Parikh VN, Caleshu C, Reuter C, Lazzeroni LC, Ingles J, Garcia J, McCaleb K, Adesiyun T, Sedaghat-Hamedani F, Kumar S, Graw S, Gigli M, Stolfo D, Dal Ferro M, Ing AY, Nussbaum R, Funke B, Wheeler MT, Hershberger RE, Cook S, Steinmetz LM, Lakdawala NK, Taylor MRG, Mestroni L, Merlo M, Sinagra G, Semsarian C, Meder B, Judge DP, Ashley E. Regional Variation in RBM20 Causes a Highly Penetrant Arrhythmogenic Cardiomyopathy. Circ Heart Fail. 2019;12:e005371.

2. Dries AM, Kirillova A, Reuter CM, Garcia J, Zouk H, Hawley M, Murray B, Tichnell C, Pilichou K, Protonotarios A, Medeiros-Domingo A, Kelly MA, Baras A, Ingles J, Semsarian C, Bauce B, Celeghin R, Basso C, Jongbloed JDH, Nussbaum RL, Funke B, Cerrone M, Mestroni L, Taylor MRG, Sinagra G, Merlo M, Saguner AM, Elliott PM, Syrris P, van Tintelen JP, James CA, Haggerty CM, Parikh VN. The genetic architecture of Plakophilin 2 cardiomyopathy. Genet Med. 2021;23:1961– 1968.

3. Anzalone AV, Randolph PB, Davis JR, Sousa AA, Koblan LW, Levy JM, Chen PJ, Wilson C, Newby GA, Raguram A, Liu DR. Search-and-replace genome editing without double-strand breaks or donor DNA. Nature. 2019;576:149–157.

4. UniProt Consortium. UniProt: a worldwide hub of protein knowledge. Nucleic Acids Res. 2019;47:D506–D515.

5. Hess GT, Frésard L, Han K, Lee CH, Li A, Cimprich KA, Montgomery SB, Bassik MC. Directed evolution using dCas9-targeted somatic hypermutation in mammalian cells. Nat Methods. 2016;13:1036–1042.

